# COVID-19 Case and Mortality Surveillance using Daily SARS-CoV-2 in Wastewater Samples adjusting for Meteorological Conditions and Sample pH

**DOI:** 10.1101/2023.07.12.23292570

**Authors:** Samantha Abelson, Johnathon Penso, Bader Alsuliman, Kristina Babler, Mark Sharkey, Mario Stevenson, George Grills, Christopher E. Mason, Helena Solo-Gabriele, Naresh Kumar

## Abstract

**Background:** Wastewater monitoring is increasingly used for community surveillance of infectious diseases, especially after the COVID-19 pandemic as the genomic footprints of pathogens shed by infected individuals can be traced in the environment. However, detection and concentration of pathogens in the environmental samples and their efficacy in predicting infectious diseases can be influenced by meteorological conditions and quality of samples.

**Objectives:** This research examines whether meteorological conditions and sample pH affect SARS-CoV-2 concentrations in wastewater samples, and whether the association of SARS-CoV-2 with COVID-19 cases and mortality improves when adjusted for meteorological conditions and sample pH value in Miami-Dade County, FL.

**Methods:** Daily wastewater samples were collected from Miami-Dade Wastewater Treatment Plant in Key Biscayne, Florida from August 2021 to August 2022. The samples were analyzed for pH and spiked with OC43. RNA was extracted from the concentrated wastewater sample and SARS-CoV-2 was quantified using qPCR. COVID-19 and mortality data were acquired from the Centers for Disease Control and Prevention (CDC) and meteorological data from the National Climatic Data Center. COVID-19 case and mortality rates were modelled with respect to time-lagged wastewater SARS-CoV-2 adjusting for meteorological conditions, and sample pH value and OC43 recovery.

**Results:** Temperature, dew point, pH values and OC43 recovery showed significant associations with wastewater SARS-CoV-2. Time-lagged wastewater SARS-CoV-2 showed significant associations with COVID-19 case and mortality incidence rates. This association improved when wastewater SARS-CoV-2 levels were adjusted for (or instrumented on) meteorological conditions, OC43 recovery, and sample pH. A 0.47% change in COVID-19 case incidence rate was associated with 1% change in wastewater SARS-CoV-2 (β ∼ 0.47; 95% CI = 0.29 – 0.64; p < 0.001). A 0.12 % change in COVID-19 mortality rate was associated with 1 % change in SARS-CoV-2 in wastewater 44 days prior. A 0.07% decline in COVID-19 mortality rate was associated with a unit increase in ambient temperature 28 days prior.

**Discussion:** Time lagged wastewater SARS-CoV-2 (and its adjustment for sample pH and RNA recovery) and meteorological conditions can be used for the surveillance of COVID-19 case and mortality. These findings can be extrapolated to improve the surveillance of other infectious diseases by proactive measurements of infectious agent(s) in the wastewater samples, adjusting for meteorological conditions and sample pH value.

## INTRODUCTION

The COVID-19 pandemic caused by the severe acute respiratory syndrome coronavirus 2 (SARS-CoV⍰2) exposed our vulnerability to the consistent threat of infectious diseases and highlighted the challenges in containing their transmission. The location- and time-specific surveillance of infectious diseases is critical for their timely containment and management. Clinical records and/or serological screening are considered as the gold standards of infectious disease surveillance. This requires large-scale testing and timely reporting of (infected) cases. However, even if clinic data are reported and updated in virtual real-time, they may not adequately capture the community level infection of the disease, because asymptomatic individuals do not seek testing and/or individuals may choose not to report their symptoms or testing results. Moreover, there is a time delay between contracting the disease and the onset of the symptoms of the disease, even though the infected person may still be shedding the virus [1]. Thus, there has been an increasing interest in alternate methods of COVID-19 surveillance, such as monitoring of the SARS-CoV-2 in wastewater and other environmental samples (Figure 1)[2]. It is well-known that individuals infected with SARS-CoV-2 shed the virus which makes its way to the environment, including wastewater. Wastewater represents the collective contributions from individuals within a given sewershed, i.e. coverage or area capturing sewer from the buildings connected to a given sewer plant. Thus, quantifying the concentration of a given pathogen in wastewater sample from the sewer plant may represent the burden of an infectious disease in a community without the community scale screening of the disease.

**Figure 1:**
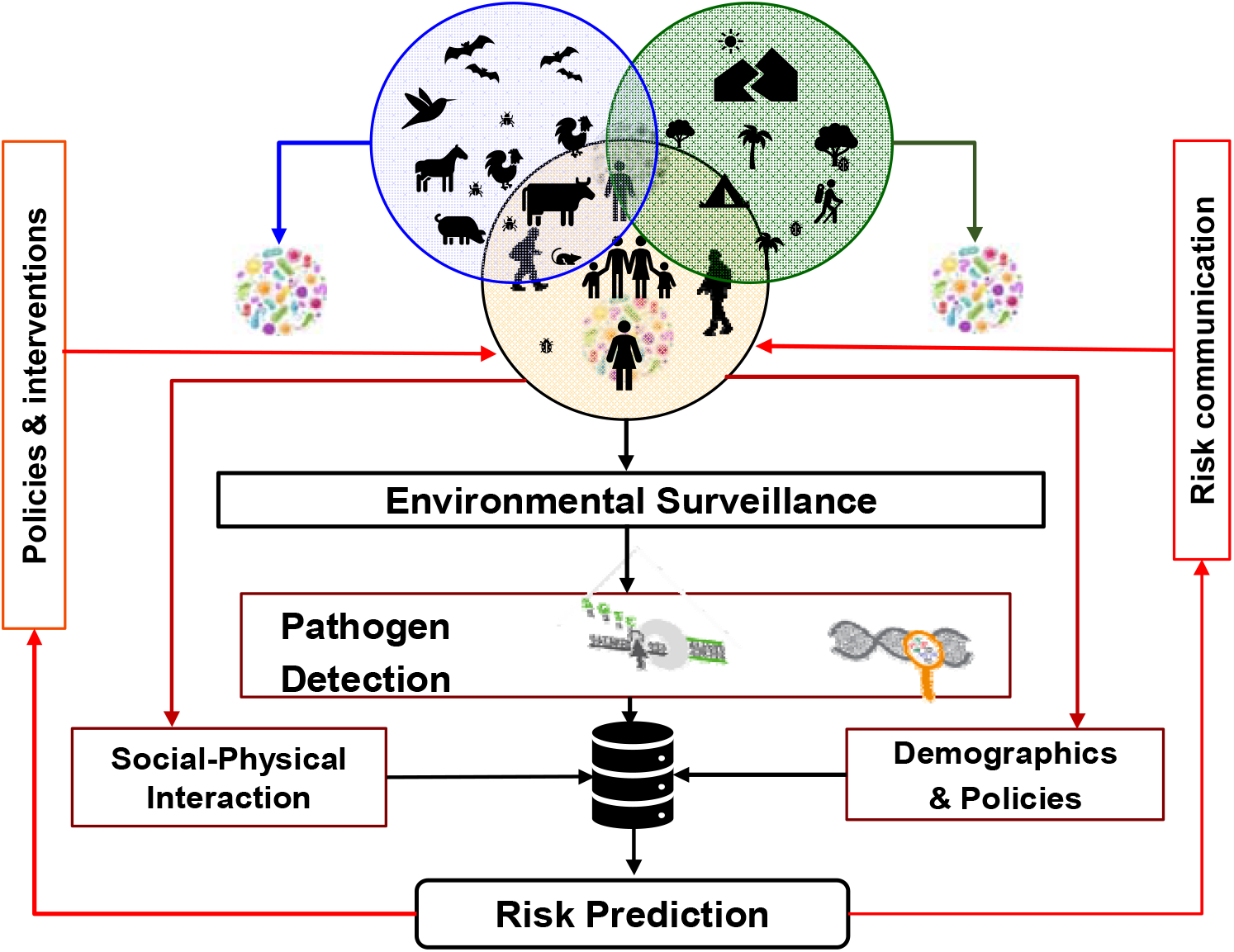
A conceptual framework of environmental surveillance of infectious agent(s) and risk prediction.

Wastewater monitoring is cost-effective and non-invasive and it can be used to detect the virus several days prior to the onset of disease symptoms. Thus, it can serve as a tool for the timely detection of COVID-19 and potentially other infectious diseases, such as influenza and monkey pox [3, 4].

Studies that utilize wastewater monitoring of SARS-COV-2 have been promising to assess community-scale infection of COVID-19 cases. Our conceptual model is based on the assumption that infected individuals (and animals) shed the virus through exhalation or their bodily fluid or feces [5], which is transferred to air, surface and wastewater (Figure 1). Thus, the genomic footprint the virus can be traced in the environmental samples, which is non-invasive [6-8]. Each type of environmental sample has benefits and limitations. However, wastewater sampling has been used extensively for assessing community scale infection of COVID-19 cases [6, 9, 10], as it offers two unique advantages. First, it represents the viral load of the individuals living in a given sewershed. Second, it is most time- and cost-effective, given one or two samples/day can provide estimate of the viral load in the community. Thus, using the time-lagged (i.e. days before clinical reporting of cases) viral load in the wastewater samples COVID-19 case and mortality prediction models can be developed, which can be cross-validated using the reported clinical data (Figure 1). Thus, viral load of SARS-CoV-2 in wastewater (or wastewater based epidemiology [WBE]) can offer early warning of COVID-19 (and potentially other infectious diseases) ranging from building to campus to country scales [6, 11, 12].

The efficacy of WBE to predict community spread of COVID-19 can be influenced by a number of factors, including sample characteristics (especially sample pH), meteorological conditions, recovery level of the virus from the sample, community scale sociodemographic characteristics and local-regional policies [13]. This research examines whether meteorological conditions, sample pH, and RNA recovery affect the concentration of SARS-CoV-2 in wastewater, and evaluates the efficacy of WBE for assessing COVID-19 cases and mortality adjusting for meteorological conditions.

## METHODS AND MATERIALS

Daily composite wastewater samples were collected from the Miami-Dade Central District Wastewater Treatment Plant (MDCWTP) located in Virginia Key, FL from August 2021 to August 2022. Each sample included the composite of hourly samples for the past 24h collected at 12:00AM (mid-night) [14]. The samples were transported to our laboratory at the University of Miami (located 30 minutes away) on ice and processed within 8h ± 1h after the collection.

Samples were thoroughly mixed and sample pH was assessed. OC43 was added to the rest of the sample as RNA recovery control. The wastewater sample was concentrated by electronegative filtration [15], and the concentrated sample was transferred to 1X DNA/RNA shield. RNA was extracted using QuickRNA-Viral Kit from Zymo Research Inc. (R1040/R1041), and eluted in 15 μL of nuclease-free water. The RNA was immediately stored in a -80ºC freezer until qPCR analysis. Volcano second Generation (V2G) reagent was used in the qPCR and a calibration curve was generated using SARS-CoV-2 synthetics RNA. Sample processing and qPCR method are detailed elsewhere [6, 9, 10]. The limit of detection of SARS-CoV-2 in wastewater samples was 100 genomic copies (GC)/L. OC43 (RNA) was also measured in the sample as a proxy of RNA recovery from the sample.

COVID-19 case and mortality incidence data from March 2020 to August 2022 were acquired from the CDC [16]. Cases and deaths were reported weekly for Miami-Dade County. COVID-19 data were modelled in two ways. First, we model COVID-19 case incidence and mortality for matching days when both data sets were available (n = 47). Second, we use interpolated daily COVID-19 case incidence and mortality rates using moving averages and observed daily SARS-CoV-2 in wastewater samples. The associations of COVID-19 case and mortality rates were explored with different time-lagged SARS-CoV-2 concentration in wastewater samples, which guided our strategy to determine the optimal lag for wastewater SARS-CoV-2 and its association with COVID-19 morbidity and mortality rates.

Ambient temperature and sample pH can influence RNA recovery and SARS-CoV-2 viability in wastewater. Thus, we examine the relationship between SARS-CoV-2 concentration in wastewater and meteorological conditions, sample pH and OC43 (as a proxy of RNA recovery). Based on these relationships, we adjust (or instrument) wastewater SARS-CoV-2. Ambient temperature and humidity are shown to influence the viability of the virus in wastewater and air samples. Temperature and humidity also affect the transmission of COVID-19 and the virulence of the disease through bronchodilation and bronchoconstriction. Thus, meteorological conditions served as both exogenous and endogenous variables in instrumental regression (see Kumar et al. for details on instrumental regression [17]). We modelled COVID-19 cases and mortality with respect to time-lagged (and moving average of) instrumented SARS-CoV-2 in wastewater samples and time-lagged ambient temperature using log-linear time-series instrumental regression (using xtivreg function in STATA ver 14.2 [18]). Time-lagged correlation between COVID-19 cases and COVID-19 mortality were also explored.

## RESULTS

Daily wastewater samples were collected for 371 days from August 2021 to August 2022. The daily concentration and frequency of SARS-CoV-2 RNA detection in wastewater varied during this time. Of the 371 days sampled, SARS-CoV-2 was detected on 271 days (73.05%) and it was below the detection limit on 100 days (26.9%). Descriptive statistics of sample characteristics, SARS-CoV-2 concentrations and meteorological conditions are presented in Table 1. The average concentration of wastewater SARS-CoV-2 varied during the study period (Figure 2). The sample pH value, ambient dew point, and RNA recovery (measured using OC43) were significantly associated with the concentration of SARs-CoV-2 in wastewater (Table 2).

**Table 1:** Descriptive statistics of wastewater sample, SARS-CoV-2 in wastewater and meteorological conditions, August 2021 to August 2023

| Variables | Daily average (95% CI in parenthesis) |
| --- | --- |
| pH value of wastewater sample collected from the plant | 7.31 (7.30- 7.33) |
| pH value of WW sample for electronegative filtration | 4.21 (4.18- 4.23) |
| SARS-CoV-2 (genomic copies [GC]/L) | 9061.93 (6732.31- 11391.54) |
| OC43 recovery (GC/L) | 6.04 (5.48- 6.60) |
| Wind direction (°; 0=N, 90E, 180 =South, 270=W | 148.94 (142.35- 155.52) |
| Wind velocity (m/s) | 42.57 (40.94- 44.20) |
| Ceiling height (m) | 14469.83 (13981.90- 14957.76) |
| Visibility (m) | 15511.31 (15409.22- 15613.40) |
| Temperature (° C) | 25.63 (25.30- 25.96) |
| Dew point (° C) | 20.24 (19.83- 20.65) |
| Sea level atmospheric pressure (mb) | 1017.38 (1017.09- 1017.67) |
| Relative humidity (%) | 74.56 (73.80- 75.32) |
| Daily precipitation (mm) | 4.80 (3.26- 6.34) |

**Table 2:** Relationship between SARS-CoV-2 in wastewater samples and meteorological conditions, pH value, and RNA recovery.

| Covariates | $\log_e(\text{SARS-CoV-2})$ | $\log_e(\text{SARS-CoV-2}^\alpha)$ |
| --- | --- | --- |
| pH | -4.250***<br>(-7.268 - -1.233) | -2.102**<br>(-3.743 - -0.460) |
| Dew point (° C) | 0.307***<br>(0.209 - 0.405) | 0.075***<br>(0.019 - 0.132) |
| $\log_e(\text{precipitation mm})$ | 0.006<br>(-0.244 - 0.257) | -0.068<br>(-0.199 - 0.063) |
| $\log_e(\% \text{ OC43 recovery})$ | 2.137***<br>(1.610 - 2.665) | 0.845***<br>(0.557 - 1.134) |
| Observations | 371 | 271 |
| R<sup>2</sup> | 0.217 | 0.141 |
| *** p<0.01, ** p<0.05, * p<0.1; 95% CI in parentheses;<br><sup>α</sup> excluding data when SARS-CoV-2 was below detection limit |  |  |

**Figure 2.**
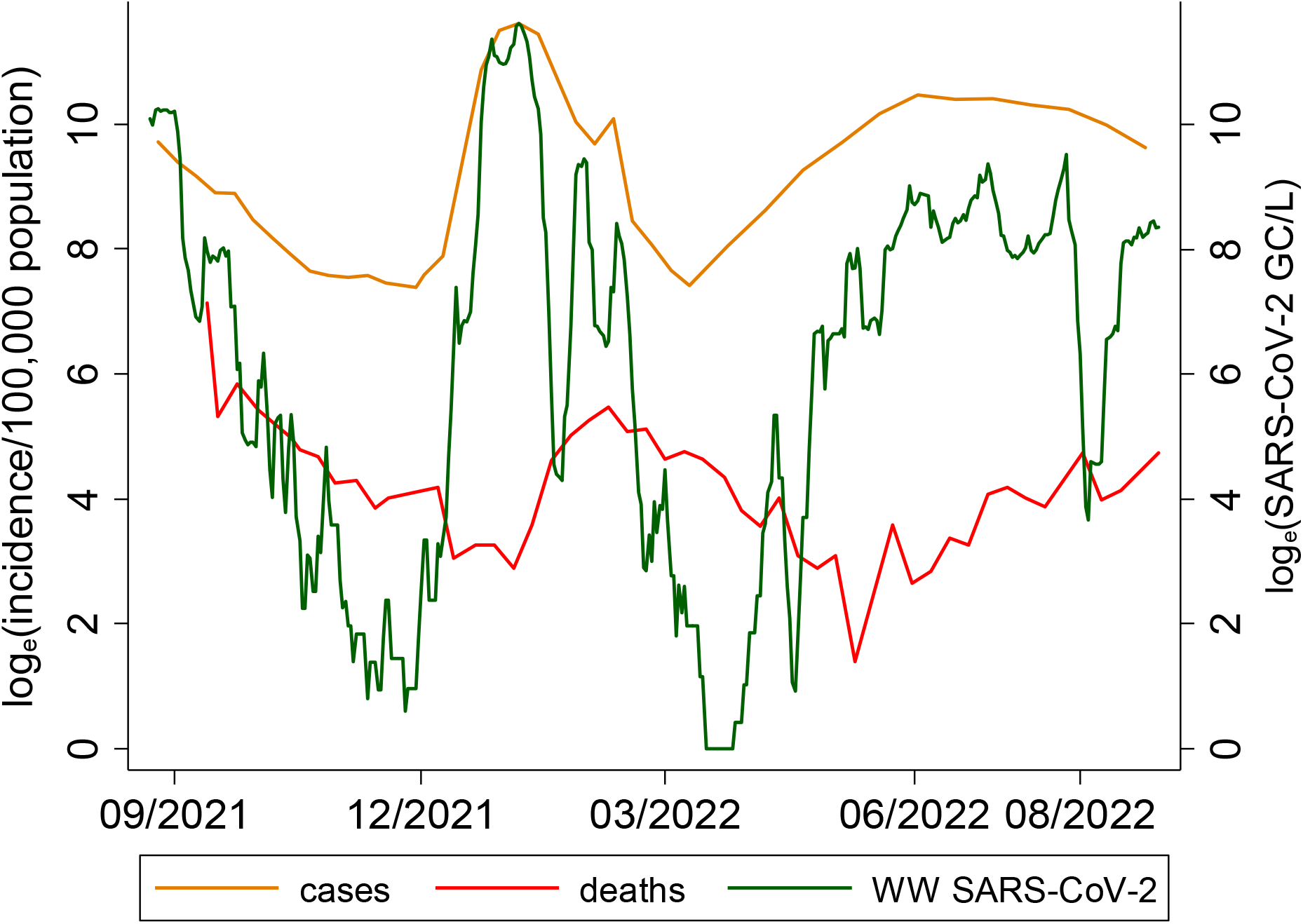
:COVID-19 cases, COVID-19 mortality in Miami-Dade County and SARS-CoV-2 in wastewater samples collected from MDC Central District wastewater plant 8/23/2021 to 8/30/2022.

A 1% decline in pH value (more acidic) was associated with a 4.25% decline in concentration of SARS-CoV-2 in wastewater (CI = -7.268 --1.233, n = 371). A 1% increase in dewpoint was associated with a 0.307% increase in the concentration of SAR-CoV-2 in wastewater (CI = 0.209 – 0.405, n = 371). Precipitation did not show significant association with the concentration of SARS-CoV-2 in wastewater. OC43 recovery (i.e. the ratio OC43 genomic copies added to the wastewater samples to OC43 genomic copies extracted from the sample), which served as a measure of RNA recovery, was strongly associated with the concentration of SARS-CoV-2. A 1% increase in OC43 recovery was associated with a 2.14% increase in the concentration of SARS-CoV-2 in wastewater (CI = 1.610 – 2.665, n = 371).

Cases and deaths were reported weekly for Miami-Dade County. Of the 53 weeks when daily wastewater samples were collected, new COVID-19 cases were reported weekly for 41 weeks, and weekly COVID-19 deaths were reported for 47 weeks. Both COVID-19 case and mortality (incidence) rates were associated with time-lagged concentration of SARS-CoV-2 in wastewater samples (Figure 2). However, the strength of the relationships of COVID-19 cases and mortality with the wastewater SARS-COV-2 varied with respect to change in lag period (Figure 3)

**Figure 3:**
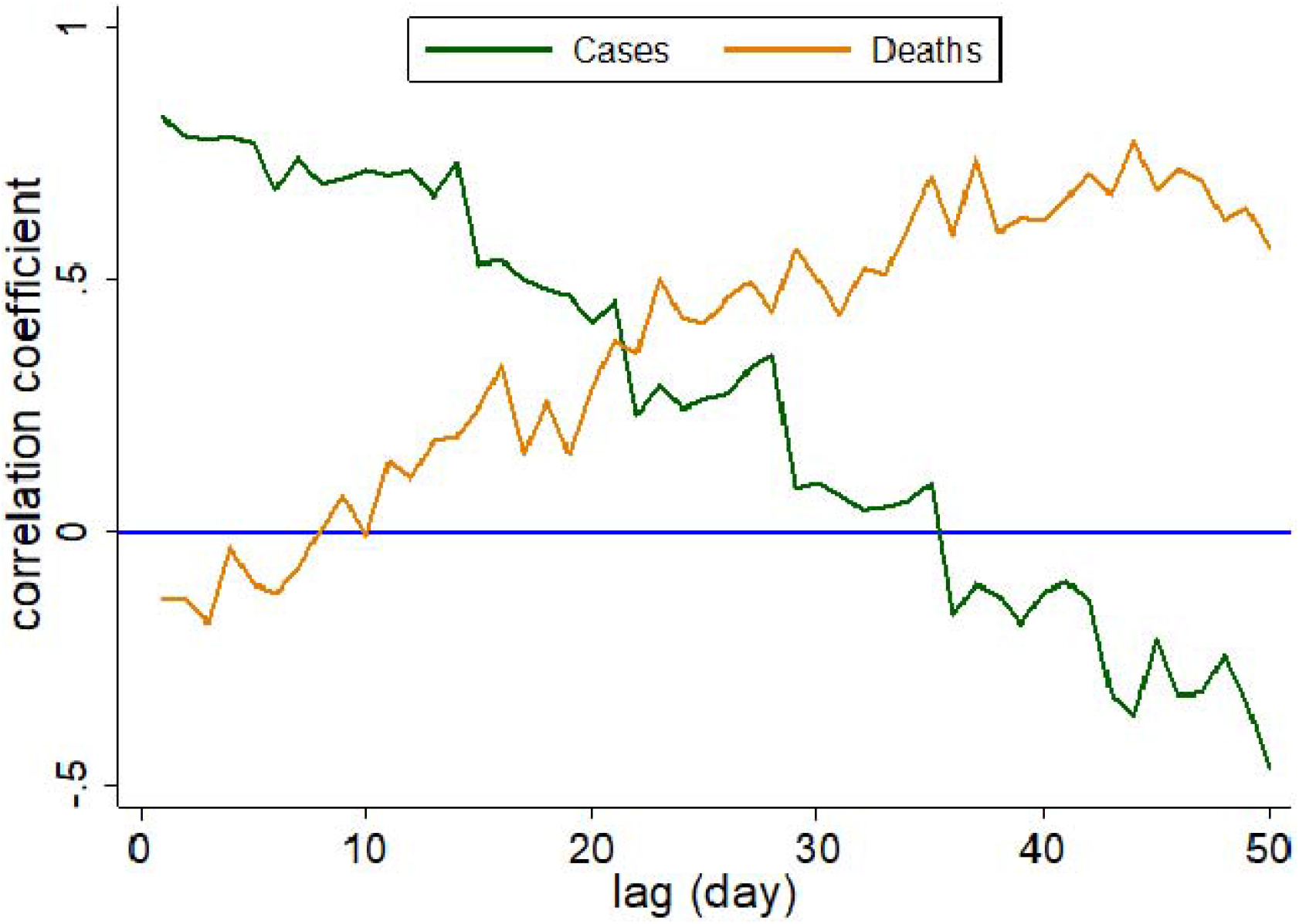
Coefficient of correlation between time-lagged SARS-CoV-2 in wastewater samples and COVID-19 cases and mortality rate in Miami-Dade, August 2021 to August 2022.

The strength of correlation between COVID-19 deaths and time-lagged SARS-CoV-2 in wastewater increased with the increase in time-lag. However, the strength of correlation between COVID-19 cases and time-lagged SARS-CoV-2 declined with the increase in lag period. For example, 1-day lagged SAR-CoV-2 showed the strongest positive association with COVID-19 cases (Figure 4a). However, this association became negative and insignificant for 37 day lagged SAR-CoV-2 (Figure 4b). COVID-19 mortality and one-day lagged wastewater SARS-CoV-2 showed a negative and insignificant relationship. However, COVID-19 mortality showed a strong positive association (ρ ∼ 0.76) with the 37-day lagged wastewater SARS-CoV-2 (Figure 4d).

**Figure 4(a-d).**
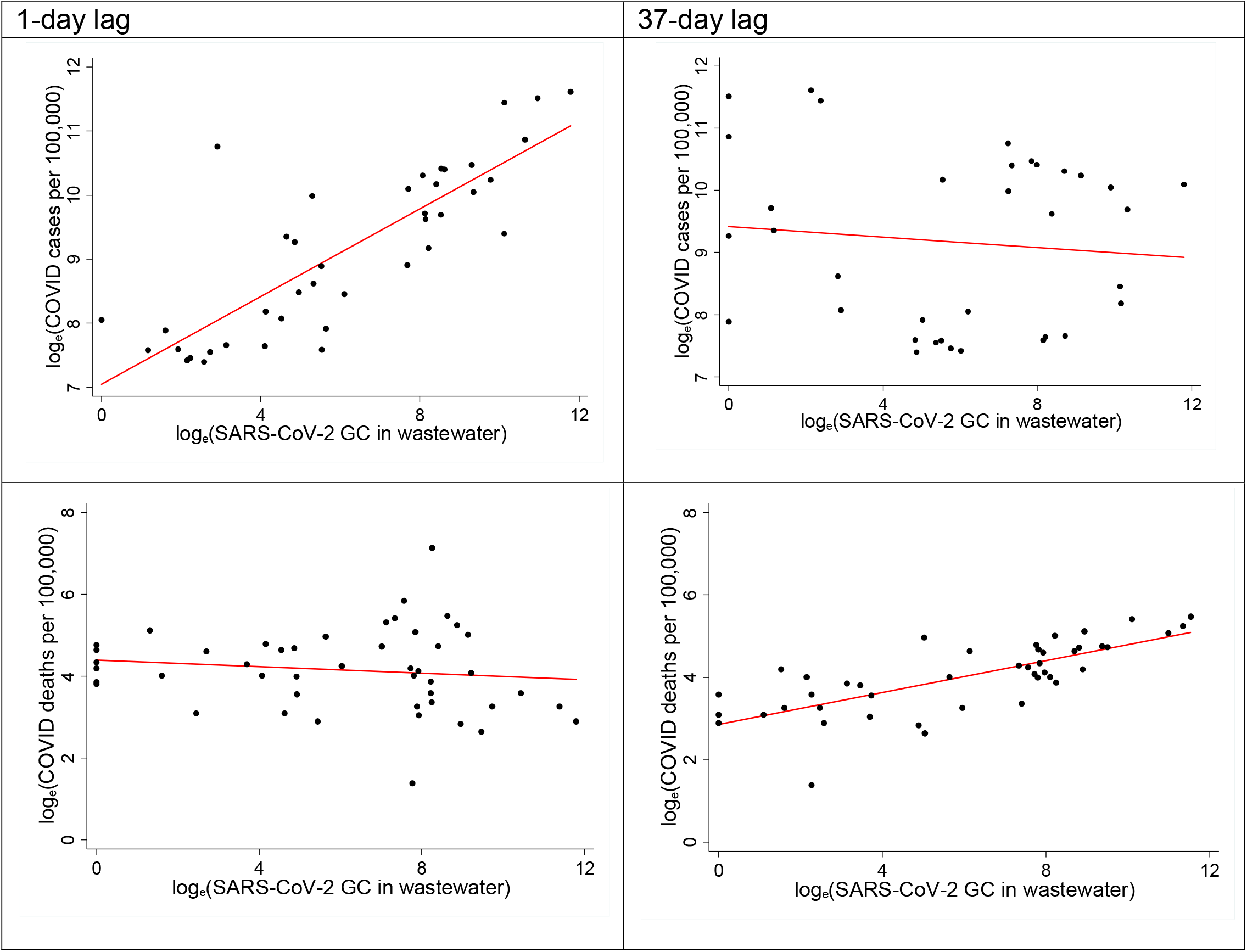
:COVID-19 case incidence and mortality with respect to time lagged SARS-CoV-2 in wastewater.

**Figure 5:**
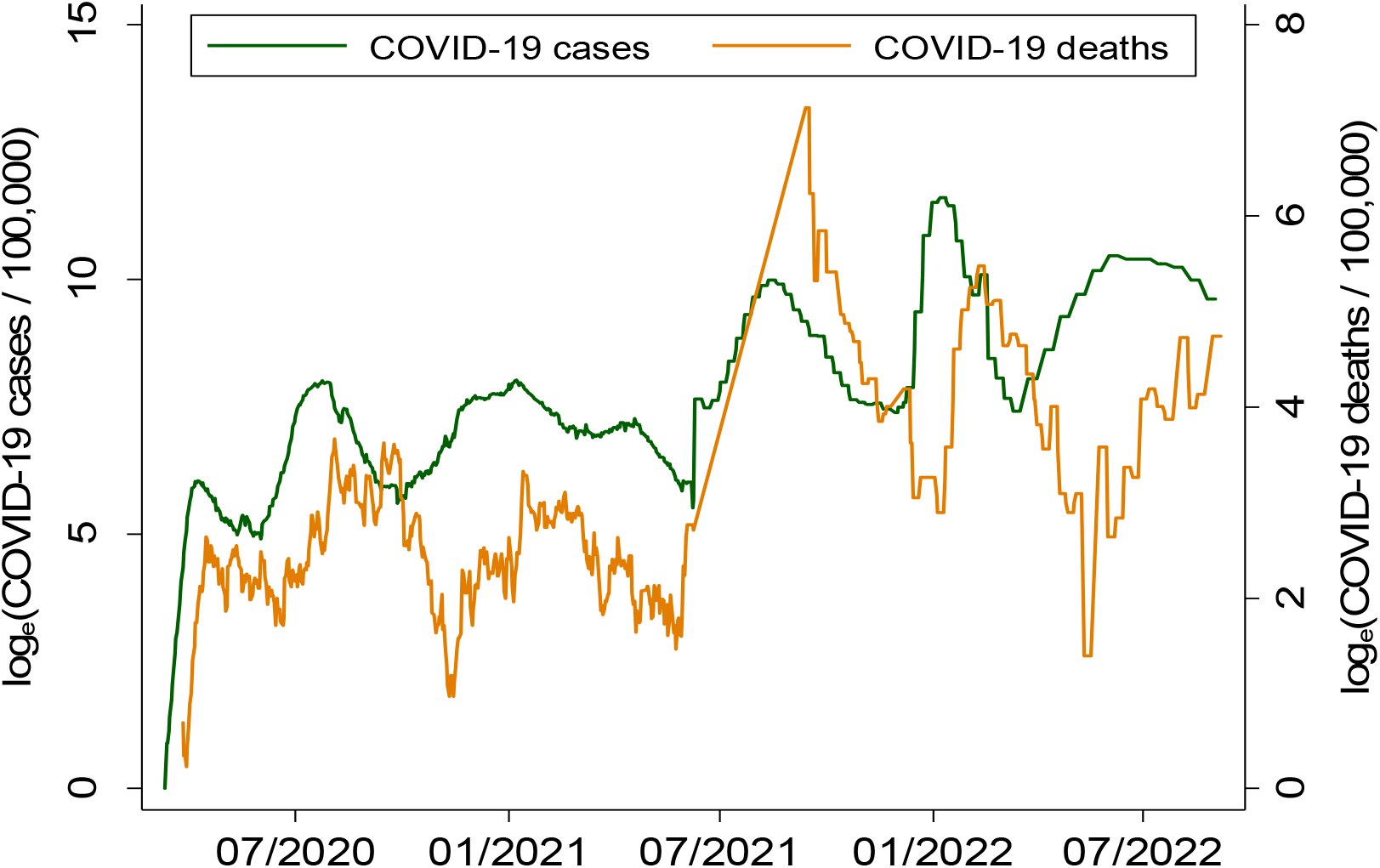
Time-lag between COVID-19 cases and COVID-19 mortality.

COVID-19 case rates and time-lagged COVID-19 mortality rates were autocorrelated. On average, the COVID-19 mortality trend lagged about 35 days behind the trends in COVID-19 cases. For example, if COVID-19 mortality peaked 35 days after the peak of COVID-19 cases. The highest peak of COVID-19 mortality was observed in September 2021 followed by another peak in January of 2022.

COVID-19 cases and mortality were modelled with respect to SARS-CoV-2 in wastewater samples adjusted for meteorology conditions and OC43 recovery (Table 3). A 0.47% change in COVID-19 case incidence rate was associated with a 1% change in SARS-CoV-2 in wastewater adjusting for OC43 recovery (β ∼ 0.47; 95% CI = 0.29 – 0.64; p < 0.001; n = 37). A 0.37% change in the seven days moving average of COVID-19 case incidence rate was associated with a 1% change in the seven days moving average of SARS-CoV-2 in wastewater (β ∼ 0.37; 95% CI = 0.33 – 0.44; p < 0.001; n = 343). A 28-day lag in ambient temperature showed the strongest association with COVID-19 case incidence rate (β ∼ -0.045; 95% CI = -0.072 – 0.019; p < 0.001; n = 343).

**Table 3:**
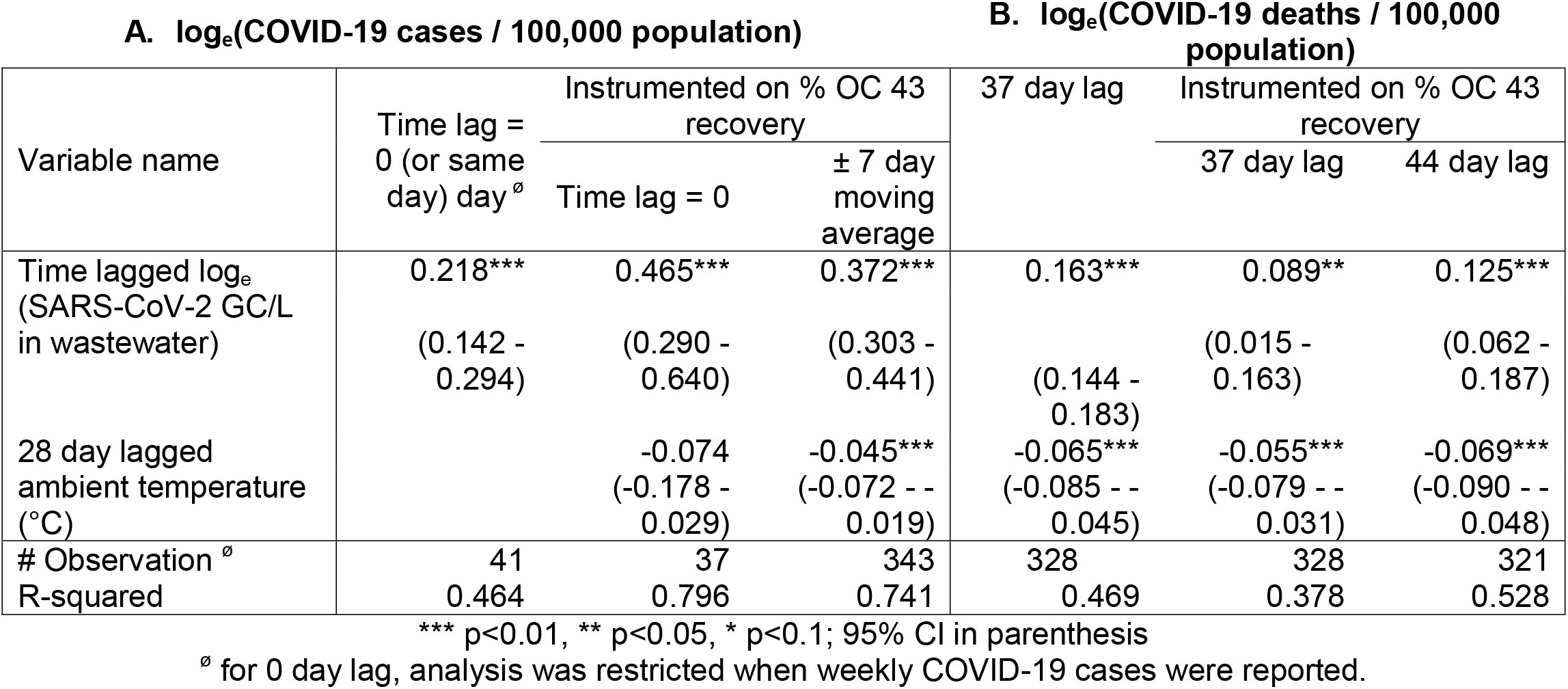
Association between SARS-CoV-2 and COVID-19 cases per 100,000 population (column A) and association between SARS-CoV-2 and COVID-19 deaths per 100,000 population (column B).

COVID-19 mortality rate showed the strongest association with the 44 lagged log_e_(SARS-CoV-2 in wastewater) (β ∼ 0.125; 95% CI = 0.062 -0.187; p < 0.001; n = 321). A 7% change in COVID-19 mortality rate was associated with a unit change in ambient temperature 28 days prior (β ∼ -0.069; 95% CI = -0.090 --0.048; p < 0.001; 321). COVID-19 case and mortality incidence rates in Miami-Dade County exhibit strong seasonal patterns, with case incidence peaked in September and mortality rate peaked in January (Figure 6a and 6b).

**Figure 6a and bb.**
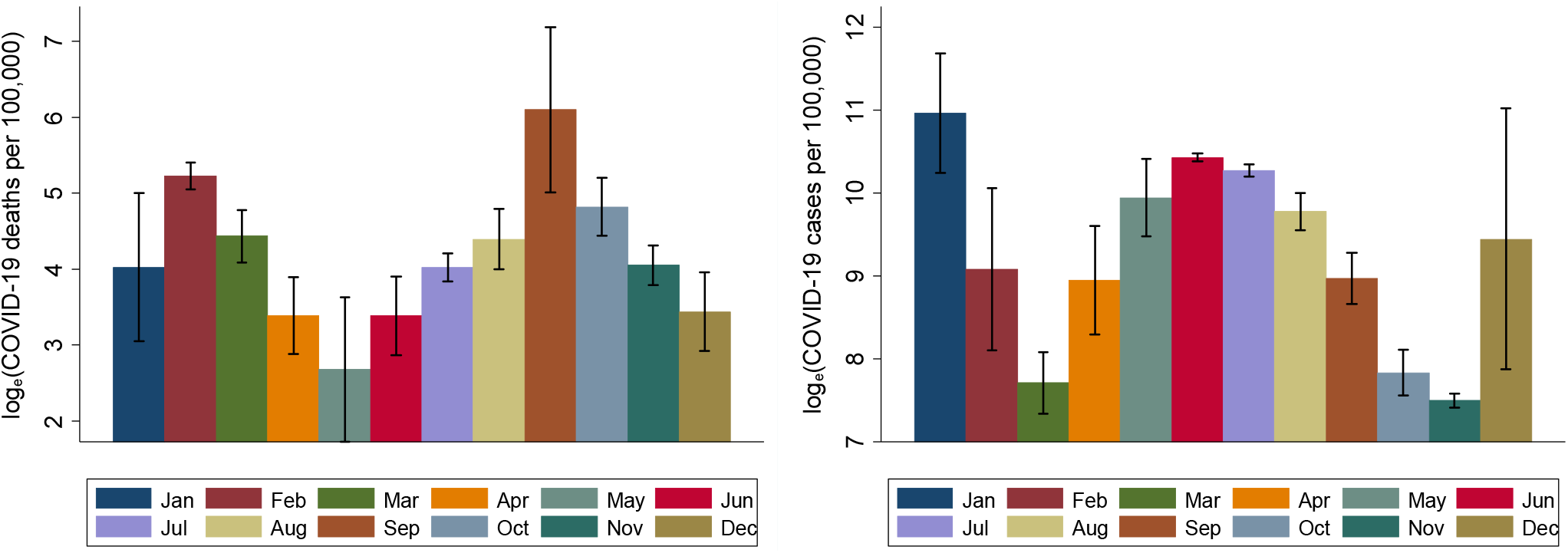
: Monthly COVID-19 case incidence (left) and mortality rate (right) in Miami-Dade, March 2020 to August 2022 based on clinical data from the CDC.

## DISCUSSION

Five findings emerge from this research. First, COVID-19 mortality trend lagged five weeks behind the trend of COVID-19 cases. Second, ambient temperature and sample pH (a measure of acidic/basic level) impacted the concentration of SARS-CoV-2 in the wastewater. Third, time-lagged wastewater SARS-CoV-2 correlated strongly with COVID-19 case and mortality rates. Fourth, time-lagged wastewater SARS-CoV-2 and time-lagged ambient temperature showed strong associations with COVID-19 case and mortality incidence rates. Finally, the efficacy of wastewater surveillance for assessing community level infection and mortality rates due to COVID-19 improved when SARS-CoV-2 concentrations were adjusted (or instrumented on) for meteorological conditions, sample pH and RNA recovery measure. Some of these findings are consistent with the emerging literature. For example, previous studies report associations between COVID-19 case rate and time lagged SARS-CoV-2 detection in environmental samples, including wastewater, and this association for COVID-19 cases between 4 to 9 days rather than 1 day as our findings suggest [19]. Previous research used wastewater and COVID-19 (reported) data from a university campus, which represented a smaller sewershed compared to the size of sewershed of present study. Thus, size of sewershed can impact the time-lag between the detection or level of pathogens in the environmental samples and clinical reporting of diseases caused by the pathogens. Our findings are consistent with those reported in other studies, which also show significant associations between meteorological conditions, particularly ambient air temperature, and SARS-CoV-2 concentration in environmental samples [20]. In our study, dew point (a proxy measure of relative humidity) and pH of the wastewater were most strongly associated with SARS-CoV-2 in the wastewater samples.

Some of the findings reported in this research make novel contributions. First, we demonstrate that wastewater SARS-CoV-2 detection and concentration can be influenced by meteorological conditions (especially dew point), sample pH value and recovery level of the (targeted) infectious agent. Thus, the paper provides an insight into how to improve the efficacy of wastewater surveillance to gauge location- and time-specific burden of infectious diseases by adjusting for meteorological conditions, pH value, and recovery level of the infectious agent in the environmental samples. Second, this research identified the lag period between the peak of COVID-19 cases and mortality, and wastewater SARS-CoV-2 and ambient temperature. Thus, the detection (and/or concentration) of an infectious agent in wastewater samples and meteorological conditions can provide early guidance on the community level infection and potential disease outbreaks. Thus, concentration and/or detection of an infectious agent in the environmental sample can offer an opportunity for early interventions to contain the transmission (or spread) of infectious diseases.

Our research shows an inverse relationship between 4-week lagged ambient temperature and COVID-19 case and mortality rates. However, the relationship between ambient temperature and COVID-19 cases and mortality has been inconsistent in the literature. Some studies suggest positive associations between ambient temperature and COVID-19 cases and mortality [21, 22]. Another study that adjusted for population mobility showed an inverse relationship between temperature and COVID-19 transmission, suggesting that an increase in ambient temperature was associated with a decrease in the incidence of COVID-19 [23].

The relationship between ambient temperature and infectious diseases needs to be interpreted with caution, because this relationship is complex and non-linear. Moreover, temperature plays direct and indirect roles in the transmission of infectious diseases. Changes in temperature directly affect biophysiological response. For example, heat exposure increases cardiovascular pressure, which in turn affects respiratory and immune systems, and responses to the infectious agent, such as COVID-19 [24]. Elevated temperature, as suggested in this research, also affect the viability of the virus through heat inactivation [25]. Other studies suggest that cool air may promote SARS-CoV-2 growth in airways. Colder outdoor temperatures may contribute to the severity of disease, as observed with other respiratory diseases, such as influenza [26]. Given the complexity of non-linear (in)direct relationship between temperature and infectious diseases, the findings of this study must be interpreted within the scope and regional domain of this research, i.e. regional sub-tropical climate as well as direct and indirect relationship as conceptualized and modelled in our research.

This research documents the relationship between time lagged SARS-CoV-2 in wastewater and COVID-19 cases and mortality rates using high resolution (daily) data for one full year. Whereas previous long-term wastewater monitoring has mostly compared data to COVID-19 case incidence only. The time lagged associations differed significantly for COVID-19 cases and COVID-19 mortality. Previous research has primarily focused on either time lagged COVID-19 cases, or time lagged COVID-19 mortality. This study distinctly shows how both COVID-19 case and mortality rates are associated with time-lagged wastewater SARS-CoV-2. Studies that examine both COVID-19 case and mortality incidence, they report 7-12 days lags between the peaks of COVID-19 cases and COVID-19 mortality [27, 28]. This paper shows longer time-lag between the peak of COVID-19 cases and the peak of COVID-19 mortality, suggesting longer hospitalizations following the diagnosis of the disease. It is important to note that previous studies used data from 2020 and 2021, when vaccine and other treatment modalities were unavailable. In our research, we also included data from 2022, when vaccine and other treatments were available, which could have delayed mortality.

Despite the novel contributions of this paper, the findings reported in the paper must be interpreted in the light of its limitations. First, the efficacy of SARS-CoV-2 in the wastewater samples to predict COVID-19 cases and mortality can be influenced by the reliability of virus recovery from the wastewater samples. For example, the detection and recovery of the virus in the sample can be subject to the physical and chemical composition of the sample and processing error. As we observed in this research, decline in pH value (meaning more acidic condition) seems to have impacted virus RNA. Second, the mismatch in the temporal (and spatial) resolution of health and environmental data can influence model inferences. For example, COVID-19 case incidence and mortality rates were reported weekly and wastewater samples were collected daily. Thus, matching both data sets by exact day resulted in reduced sample size and interpolating weekly data by every day introduced uncertainty. Third, the findings reported in this research did not account for confounders, such as socio-demographic characteristics or pre-existing health conditions of individuals, vaccination, COVID-19 control measures (mask use and quarantine). Because these data were unavailable at higher spatiotemporal scale. Finally, our models lacked adjustment for the existing COVID-19 cases or a measure of infectious disease diffusion and transmission.

## CONCLUSIONS

This paper has important implications for infectious disease surveillance by emphasizing that wastewater surveillance can serve as a non-invasive tool for the early detection and prediction of infectious diseases (outbreak) at a community scale. When adjusted for RNA recovery and time-lagged meteorological conditions, our model explained 80% of the total variance in COVID-19 cases. Our model that included time-lagged ambient temperature and time lagged SARS-CoV-2 explained 52% of the total variance in COVID-19 mortality rate at a community scale. Therefore, wastewater surveillance of other infectious agents, including the RNA/DNA of common respiratory viruses, such as influenza A and B and RSV, can be used for assessing community level infection of such diseases when adjusted for the recovery level of the infectious agent and meteorological conditions. Thus, these factors must be considered in modeling the prediction of infectious diseases with the aid of environmental surveillance of the infectious agent.

In summary, genomic profiling of environmental samples across-space and time can shed light on the spatiotemporal dynamics of infectious disease(s) in a community. This, in turn, has potential to guide public health interventions and risk communications strategies for minimizing the transmission of disease and protecting public health. Wastewater represents one of the environmental media for detecting infectious diseases. Future efforts should also include genomic profiling of other environmental media (aerosols and surfaces) at different spatiotemporal scales to understand the spatiotemporal dynamics of disease transmission needed to develop location- and time-specific infectious disease risk surveillance system for targeted location- and time-specific public health interventions.

## Data Availability

All data produced in the present study are available upon reasonable request to the authors

## Acknowledgements

We are grateful to the staff members of Miami-Dade Wastewater Treatment Plant for allowing us to collect daily wastewater samples. This work in part was supported by NIH (U01DA053941). All clinical data (COVID-19 case and mortality rates for Miami-Dade County) are in public domain and daily SARS-CoV-2 concentration in the wastewater samples can be provided upon a reasonable request to the corresponding author.

